# Oral Cancer Prevalence, Mortality, and Costs in Medicaid and Commercial Insurance Claims Data

**DOI:** 10.1101/2022.04.25.22274259

**Authors:** Eric P. Tranby, Lisa J. Heaton, Scott Tomar, Abigail L. Kelly, Gulielma Leonard Fager, Mary Backley, Julie Frantsve-Hawley

**Affiliations:** Analysis and Evaluation, CareQuest Institute for Oral Health, 465 Medford Street Boston, MA 02129; Department of Pediatric Dentistry, College of Dentistry, University of Illinois Chicago, 801 South Paulina Street, IL 60612; PPD, Inc. 3900 Paramount Parkway, Morrisville, NC 27560; Maryland Dental Action Coalition, 10015 Old Columbia Road, Suite B-215, Columbia, MD 21046; The Aspen Group (TAG) Oral Care Center for Excellence, 800 W. Fulton Market, Chicago, IL 60607

**Keywords:** Oral Cancer, Oropharyngeal Cancer, Incidence, Prevalence, Health Care Costs

## Abstract

**Objectives:** This study compared prevalence, incidence, mortality rates, treatment costs, and risk factors for oral and oropharyngeal cancer (OC/OPC) between two large cohorts of adults in 2012–2019.

**Methods:** Medicaid and commercial claims data were from the IBM Watson Health MarketScan Database. Logistic regression analyses estimated incidence and risk factors for OC/OPC. Mortality was calculated by merging deceased individuals’ Medicaid files with those of the existing cancer cohort. Costs were calculated by summing costs of outpatient and inpatient services.

**Results:** The prevalence of OC/OPC in the Medicaid cohort decreased each year (129.8 cases per 100,000 enrollees in 2012 to 88.5 in 2019); commercial enrollees showed a lower and more stable prevalence (64.7 per 100,000 in 2012 and 2019). Incidence trended downward in both cohorts, with higher incidence in the Medicaid (51.4–37.6 cases per 100,000) than in the commercial cohort (31.9–31.0 per 100,000). OC/OPC mortality rates decreased in the Medicaid cohort during 2012–2014 but increased in the commercial cohort. Total OC/OPC treatment costs were higher for commercial enrollees by an average of $8.6 million during 2016–2019. In both cohorts, incidence of OC/OPC was higher among adults who were older, male, white, used tobacco or alcohol, or had prior HIV/AIDS diagnosis, and lower among those who had seen a dentist within the prior year.

**Conclusions:** Medicaid enrollees experienced higher OC/OPC incidence, prevalence, and mortality compared with commercially insured adults. Having seen a dentist within the prior year was associated with a lower risk of OC/OPC diagnosis.

## Introduction

Oral cancer is the sixth most common cancer worldwide,^(1-3)^, accounting globally for 377,713 new cases and 177,757 deaths in 2020,^(4)^ representing an increase in new cases from 2018 (354,864 new cases, 177,384 deaths in 2018).^(5)^ In the United States (U.S.), the American Cancer Society estimates that approximately 54,010 new cases of cancer in the oral cavity or oropharyngeal region will be diagnosed in 2021 (about 3% of the cancers diagnosed in the U.S. annually at a per-incidence cost of $45,000-$70,000 ^(6, 7)^), and about 10,850 people will die from OC/OPC this year.^(8)^ The average 5-year survival rate in the U.S. is 64.3%, but this is stage-dependent. Approximately 70% of cases are diagnosed at a late stage, reducing the 5-year survival rate from 83.7% when diagnosis occurs at a localized state to 38.5% if the cancer has metastasized when diagnosed.^(9)^

Although the term “oral cancer” is used commonly to describe cancers that occur in the oral cavity and the oropharyngeal area, it is useful to distinguish between locations because cancers in different sites are linked with different risk factors. Conway and colleagues describe *oral cavity cancer* sites as including the inner lip, parts of the tongue apart from the base and lingual tonsil, gingiva, floor of the mouth, palate and “other unspecified parts of the mouth.”^(10)^ Meanwhile, *oropharyngeal cancer* occurs at sites including the base of the tongue, the lingual tonsil and tonsil, the oropharynx, and the pharynx including Waldeyer’s ring.^(3, 10, 11)^ For the purposes of this paper, the term oral and oropharyngeal cancer (OC/OPC) will be used unless otherwise specified.

The incidence rates of cancer in some areas of the mouth (lip, gingiva, floor of the mouth) have been decreasing in the U.S. over the last few decades, which has been attributed to decreasing rates of tobacco use and alcohol consumption overall.^(6, 9)^ Meanwhile, the incidence of oropharyngeal cancers in the base of the tongue, pharyngeal wall, tonsil, and soft palate has increased, which some have associated with =an increase in human papillomavirus (HPV).^(1, 9, 10, 12-17)^ In a sample of over 5,000 head and neck squamous cell carcinoma samples, Kreimer and colleagues noted that HPV was found in more oropharyngeal cancer samples (35.6%) than oral cancer samples (23.5%).^(18)^ While age-adjusted incidence rates of oral squamous cell carcinoma (OSSC) for most oral sites (e.g., lip, gingiva, and floor of the mouth) have decreased over the last few decades, incidence rates for cancers of the anterior two-thirds of the tongue and oropharyngeal sites (e.g., posterior third of the tongue, pharyngeal wall, tonsil and soft palate) have increased over the same time.^(9, 19, 20)^

The incidence of oral cancer is 2-3 times higher in men than women.^(1, 10, 12)^ However, there has been a surprising increase in tongue cancer in younger women without a history of tobacco or alcohol use, although whether this is due to a unique presentation of oral cancer in this population or other unknown factors has yet to be determined.^(12, 19, 21, 22)^ Common risk factors for oral and oropharyngeal cancer are tobacco use,^(1, 10, 23, 24)^ alcohol consumption,^(2, 10)^ older age,^(3, 25)^ human immunodeficiency virus (HIV),^(26)^ and HPV infection.^(27-29)^

In 2019, the American Dental Association recommended that “dentists conduct routine visual and tactile examinations for oral and oropharyngeal cancer for all patients”,^(30)^ recognizing the key role dental providers play in the early detection of OC/OPC through routine screenings.^(31, 32)^ Individuals with infrequent dental visits are often diagnosed with OC/OPC at later stages than are individuals who visit a dentist more regularly.^(33, 34)^ Along with caries and periodontal disease, OC/OPC is an oral disease with implications for overall health,^(35-37)^ emphasizing the important role dentists play in detecting and helping prevent the negative outcomes of this consequential disease.

The current study compared the prevalence, incidence, and mortality rates of OC/OPC between two large data cohorts of adults: Medicaid enrollees and individuals with commercial medical insurance. Trends across time (2012-2019), across age groups, and between sexes were also calculated, as well as the cost of OC/OPC treatment for both groups. Analyses further examined risk factors for OC/OPC in both cohorts.

## Methods

### Study Population

This study compared data from two sources included in the national IBM Watson Health MarketScan Database. The first source was Medicaid claims data from 13 de-identified states collected from 2012 through 2019 (N= 36,749,894). The second group involved data from commercial dental claims from 2012 through 2019 (N=37,542,904). Medicaid enrollees were adults aged 21 years or older who answered “yes” to having Medicaid coverage at any point during the years of 2012 through 2019. Similarly, commercially insured respondents were adults aged 21 years or older who reported “yes” to having private insurance at any point during the years of 2012 through 2019.

Both datasets were restricted to adults aged 21 years or older. In addition, data from adults aged 65 years or older were excluded, because those adults in the Medicaid database were dually eligible for medical coverage through Medicaid and Medicare, resulting in incomplete medical claims data for this age group. It was therefore decided to exclude data for individuals from this older group for both datasets. Additionally, data on race and ethnicity were not available in the commercial database.

### Data Procedures

Data on prevalence (the presence of any OC/OPC diagnosis during the given year) and incidence (the presence of a new cancer diagnosis within the given year with no prior OC/OPC diagnosis within the prior 3 years) of OC/OPC were collected from both datasets. Cases were identified by using the International Classification of Disease (ICD-10) code C000–C148, “*Malignant neoplasms of lip, oral cavity and pharynx*”. ICD-10 codes were used to further distinguish oral (C00-lip; C02-other/unspecified parts of tongue; C03-gingiva; C04-floor of mouth; C05-palate; C06- other/unspecified parts of mouth; C07-parotid gland; C08-other/unspecified major salivary glands) from oropharyngeal cancers (C01-base of tongue; C09-tonsil; C10-oropharynx; C14-other/ill-defined sites in the lip, oral cavity and pharynx).

To estimate the incidence of OC/OPC, logistic regression analyses with diagnosis of OC/OPC as the outcome were conducted for the Medicaid and commercial claims data separately. Models included the covariables of age, sex, race (Medicaid only), Elixhauser comorbidities associated with OC/OPC including tobacco use, alcohol use, a diagnosis of HIV/AIDS, and whether the enrollee had a dental visit in the prior year.

Information on mortality from OC/OPC was collected from both cohorts by pulling in all discharge statuses from inpatient data that indicated the patient had “expired” or “died” (i.e., codes 20, 41, 42, or 43). After verifying there was only one code per individual, the percentage of individuals who died from OC/OPC was calculated by merging deceased individuals’ files with those of the existing cancer cohort.

For commercial data, beginning in 2016, values indicating death or transfer to law enforcement were no longer used in the commercial claims data in to protect patients’ privacy. Therefore, comparisons in mortality rates between the two datasets were limited to 2012 through 2015, and trends were calculated for the Medicaid data for 2015 through 2019.

Costs associated with OC/OPC were calculated by summing the costs of services in the presence of an OC/OPC diagnostic code in both outpatient and inpatient data. Additionally, for patients with an OC/OPC diagnostic code each year, the costs for chemotherapy (therapy class 316) were included in the total costs (Table 2). The median cost per individual was calculated by summing treatment and chemotherapy costs per individual and then calculating the median of the total.

### Statistical Methods

Descriptive statistics were used to analyze differences in prevalence and incidence between adults covered by Medicaid or commercial insurance overall and by age group and sex. Multiple logistic regression models were used to predict incidence of OC/OPC among Medicaid and commercial insurance enrollees. Covariables included age, sex, race (Medicaid only), presence of a dental visit in the prior year, and Elixhauser comorbidities associated with OC/OPC including: tobacco use (created based on the presence of International Classification of Disease, 10th revision (ICD-10) codes ‘Z720’ (tobacco use) or ‘F17’ (nicotine dependence)), alcohol use, and diagnosis of HIV/Acquired Immune Deficiency Syndrome (HIV/AIDS). Odds ratios and 95% Wald confidence intervals were presented for significant predictors. All analyses were performed by using SAS Enterprise Guide version 7.1.

The data generated in this study are available upon request from the corresponding author.

## Results

### Demographics

Claims data from both cohorts were obtained for the period of 2012-2019. Most of the Medicaid OC/OPC sample (N=37,728) was aged 51-60 (53.8%) or 61-64 years (21.2%). About two-thirds (66.5%) of the sample were male, and 65.2% were white (Table 1). Nearly half the commercial OC/OPC cohort (N=27,166) sample were aged 51-60 years (48.9%), and 26.0% were aged 61-64 years. Approximately two-thirds of the commercial OC/OPC cohort were male (67.9%).

**Table 1:** Percentage distribution of adults in the Medicaid and commercial insurance cohorts, by demographic characteristics

|  | **Medicaid**<br>N=37,728 | **Commercial**<br>N=27,166 |
| --- | --- | --- |
| **Gender** |  |  |
| Male | 66.5 | 67.9 |
| Female | 33.5 | 32.1 |
| **Age** |  |  |
| 21-30 | 2.5 | 2.4 |
| 31-40 | 5.2 | 5.9 |
| 41-50 | 17.4 | 16.7 |
| 51-60 | 53.8% | 48.9 |
| 61-64 | 21.2 | 26.0 |
| **Race** |  |  |
| White | 65.2 | -- |
| Black | 27.7 | -- |
| Hispanic | 1.3 | -- |
| Other | 5.8 | -- |

*\*Data on race or ethnicity not available in the commercial insurance dataset*

**Table 2:** Logistic regression estimate of oral cancer incidence among Medicaid and commercial enrollees

|  | Medicaid |  |  |  | Commercial |  |  |  |
| --- | --- | --- | --- | --- | --- | --- | --- | --- |
| Effect | OR | 95% Wald Confidence Limits |  | P-value | OR | 95% Wald Confidence Limits |  | P-value |
| Age Group (ref= 21-30) |  |  |  | <0.0001 |  |  |  | <0.0001 |
| 31-40 | 1.84 | 1.36 | 2.48 |  | 2.10 | 1.41 | 3.13 |  |
| 41-50 | 5.22 | 3.97 | 6.85 |  | 4.56 | 3.19 | 6.52 |  |
| 51-60 | 13.37 | 10.36 | 17.27 |  | 11.82 | 8.44 | 16.56 |  |
| 61-64 | 17.10 | 13.12 | 22.29 |  | 17.34 | 12.31 | 24.43 |  |
| Sex (ref=male) |  |  |  | <0.0001 |  |  |  | <0.0001 |
| Female | 0.46 | 0.41 | 0.50 |  | 0.46 | 0.41 | 0.52 |  |
| Race (ref = white) |  |  |  | <0.0001 |  |  |  | -- |
| Black | 0.66 | 0.59 | 0.74 |  | -- | -- | -- | -- |
| Hispanic | 0.78 | 0.52 | 1.15 |  | -- | -- | -- | -- |
| Other | 0.84 | 0.58 | 1.21 |  | -- | -- | -- | -- |
| Elixhauser Comorbidities |  |  |  |  |  |  |  |  |
| Tobacco | 2.84 | 2.57 | 3.13 | <0.0001 | 2.21 | 1.85 | 2.65 | <0.0001 |
| Alcohol | 2.24 | 1.98 | 2.54 | <0.0001 | 2.73 | 2.03 | 3.67 | <0.0001 |
| HIV/AIDS | 1.89 | 1.38 | 2.60 | <0.0001 | 2.26 | 1.07 | 4.76 | 0.0302 |
| Dental Prior Year (ref=no) |  |  |  | <0.0001 |  |  |  | <0.0001 |
| Yes | 0.71 | 0.62 | 0.80 |  | 0.41 | 0.36 | 0.46 |  |

### Prevalence of Oral Cancer

Between 2012 and 2019, the prevalence of OC/OPC in the Medicaid claims cohort gradually decreased each year, from 129.8 cases per 100,000 enrollees in 2012 to 88.5 cases per 100,000 in 2019 (Figure 1). The commercial cohort showed a lower but more stable prevalence across the same period, with prevalence ranging between 69.9 and 64.8 per 100,000 between 2012 and 2016, then dropping slightly to 64.0, 62.1 and 64.7 in 2017, 2018, and 2019, respectively. The prevalence of oropharyngeal cancer across all years was slightly higher in both cohorts (54.4% for Medicaid and 58.7% for commercial insurance) than oral cancer (45.5% for Medicaid and 40.2% for commercial insurance). Because the prevalence estimates were similar for oral and oropharyngeal cancer (Appendix A), results are presented for both types of cancer combined going forward.

**Figure 1:**
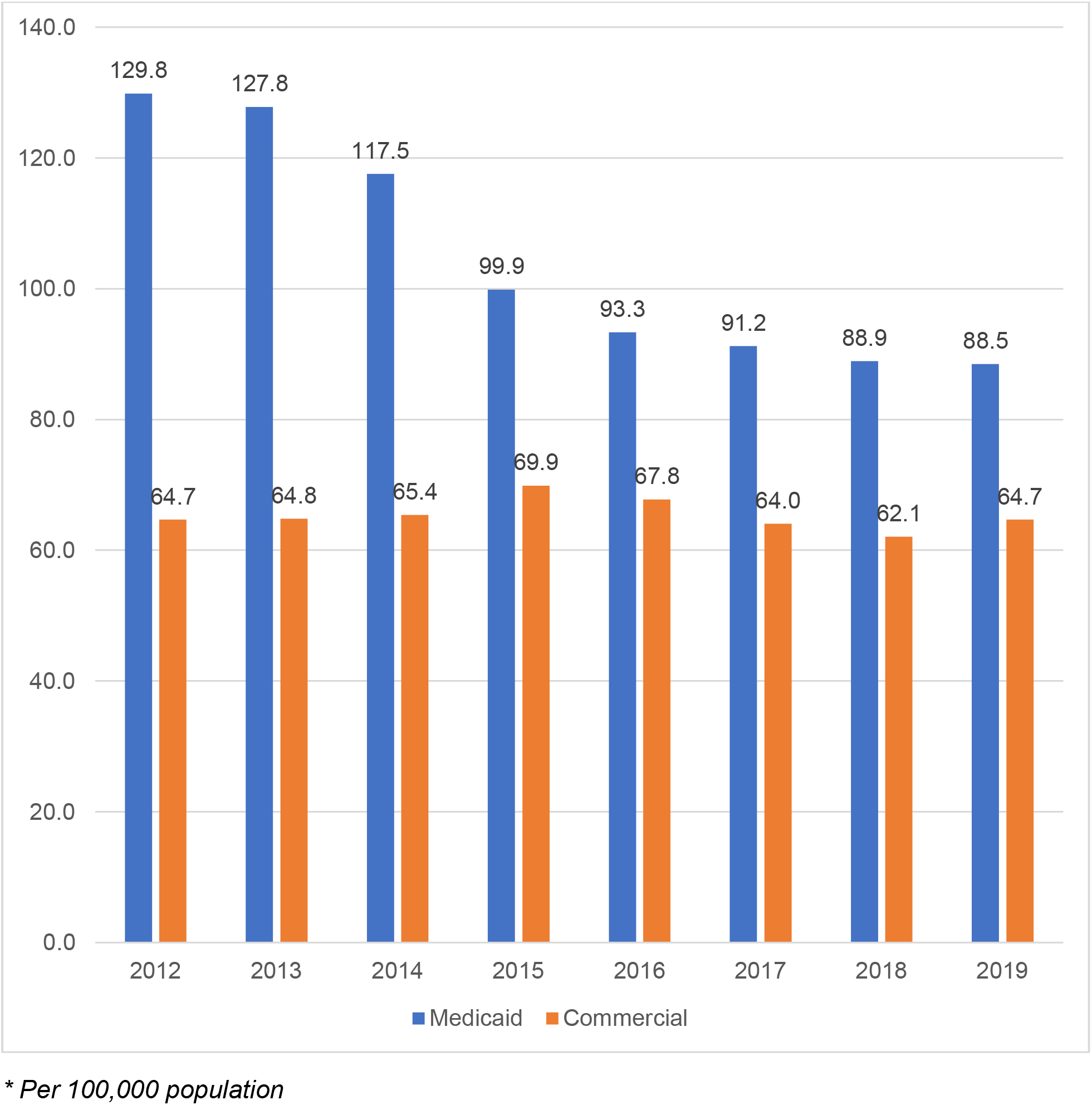
Prevalence* of oral/oropharyngeal cancer among Medicaid and commercial insurance cohorts, by year, 2012–2019.

In both cohorts, prevalence of OC/OPC increased with age (Appendix B), as did the disparity between the groups. In individuals aged 21-29 years, the prevalence was similar in both groups, although slightly higher in the Medicaid data (7.1 per 100,000 among Medicaid enrollees versus 5.8 per 100,000 among the commercial insurance enrollees). By age 61-64, the prevalence of OC/OPC among Medicaid enrollees was nearly 1.6 times higher than that of individuals with commercial insurance (325.4 versus 209.4 per 100,000, respectively).

In 2019, the prevalence was more than 3.7 times higher among males than among females in the Medicaid cohort (168.4 versus 45.5 per 100,000, respectively). While prevalence was lower among the commercial insurance cohort than among the Medicaid cohort, OC/OPC was still more prevalent among men than among women, 74.6 and 31.5 cases per 100,000, respectively.

In the Medicaid cohort in 2019, white adults experienced a higher prevalence of OC/OPC (104.1 per 100,000) compared with Black adults (60.1 per 100,000), Hispanic adults (37.4), or individuals in “other” racial categories (14.3 per 100,000).

### Incidence of Oral Cancer

The incidence of new OC/OPC cases across years was similar in both the Medicaid (23.5 oral cancer and 19.3 oropharyngeal per 100,000) and commercial datasets (18.5 oral cancer and 18.4 oropharyngeal cases per 100,000; Appendix A). The incidence rates trended downward in the Medicaid data from 2015 (51.4 cases per 100,000) through 2019 (37.6 per 100,000; Figure 2). The commercial data showed a lower overall incidence rate compared with the Medicaid data with a slight decline over time, beginning with 31.9 cases per 100,000 in 2015 and decreasing to 31.0 per 100,000 in 2019.

**Figure 2:**
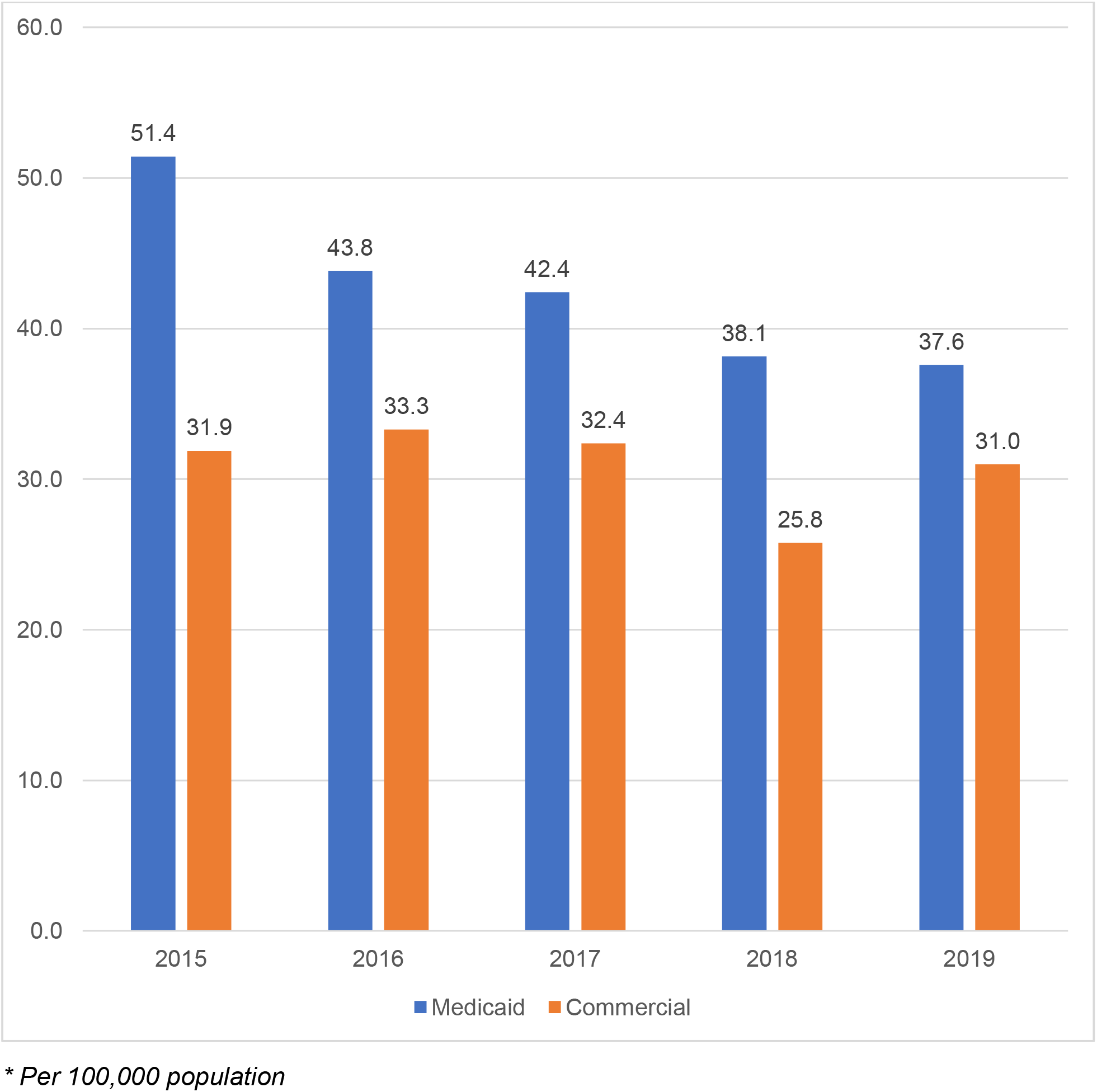
Annual incidence* of oral/oropharyngeal cancer among Medicaid and commercial insurance cohorts, by year, 2015–2019.

As with the prevalence data, the incidence of OC/OPC increased with age in both datasets (Appendix C). Across all age groups, the incidence was higher in Medicaid enrollees than in those with commercial insurance. Beginning in the 21–30 age group, prevalence of OC/OPC was the same for those with Medicaid as for those with commercial insurance (5.1 per 100,000). As age increased, the difference in incidence between the two insurance types grew. By age 61–64, the incidence of OC/OPC in Medicaid enrollees was nearly 1.6 times higher than that of individuals with commercial insurance (139.5 versus 88.2 per 100,000, respectively).

Males had a higher incidence of OC/OPC than females in both datasets. In the Medicaid cohort, male Medicaid enrollees had a higher incidence than males with commercial insurance (78.1 per 100,000 and 36.9 per 100,000 respectively). For females, the incidence was similar between datasets, with 23.7 cases per 100,000 for Medicaid enrollees and 21.3 cases for individuals with commercial insurance.

Among Medicaid enrollees, risk of OC/OPC increased with age (Table 2). Individuals aged 61–64 years had an odds ratio (OR) of 17.1 (95% CI=13.12–22.29; p<.0001) when compared to the reference group of individuals aged 21–30 years. Medicaid enrollees who had a dental visit within the prior year were less likely than those who did not to be diagnosed with OC/OPC (OR=0.71, 95% CI=0.62–0.80; p<.0001). Females were less likely than males to be diagnosed with OC/OPC (OR=0.46; 95% CI=0.41–0.50; p<.0001). White individuals were significantly less likely than Black individuals to be diagnosed (OR= 0.66; 95% CI=0.59-0.74; p<.0001). Tobacco use (OR=2.84; 95% CI=2.57–3.13; p<.0001), alcohol use (OR=2.24; 95% CI=1.98–2.54; p<.0001), and a diagnosis of HIV/AIDS (OR=1.89; 95% CI=1.38–2.60; p<.0001) were all significant predictors of an OC/OPC diagnosis.

Similar to the incidence pattern observed in the Medicaid cohort, in the commercial insurance cohort (Table 2), individuals aged 61–64 years were significantly more likely than individuals aged 21–30 years to be diagnosed (OR=17.34; 95% CI=12.31–24.43; p<.0001). Commercial insurance enrollees who had a dental visit within the prior year were less likely to be diagnosed with OC/OPC than those who had not (OR=0.41, 95% CI=0.36–0.46; p<.0001). Females were much less likely than males to be diagnosed (OR=0.46; 95% CI=0.41–0.52; p<.0001). As in the Medicaid cohort, tobacco use (OR=2.21; 95% CI=1.85–2.65; p<.0001), alcohol use (OR=2.73; 95% CI=2.03–3.67; p<.0001), and an HIV/AIDS diagnosis (OR=2.26; 95% CI=1.07–4.76; p<.0001) were all significant predictors of an OC/OPC diagnosis.

### Oral Cancer Mortality Rates

Between 2012 and 2014, OC/OPC annual mortality rates in the Medicaid cohort decreased while rates among the commercial insurance cohort increased (Figure 3). In 2012, the mortality rate was 2.03% for Medicaid enrollees and 0.95% for commercial enrollees. By 2014, the gap between the two groups had narrowed slightly, showing a mortality rate of 1.92% and 1.07% for Medicaid and commercial enrollees, respectively. Beginning in 2016, there was a leveling off in mortality rate for Medicaid enrollees across the next four years, from 1.89% in 2016 to 1.86% in 2019.

**Figure 3:**
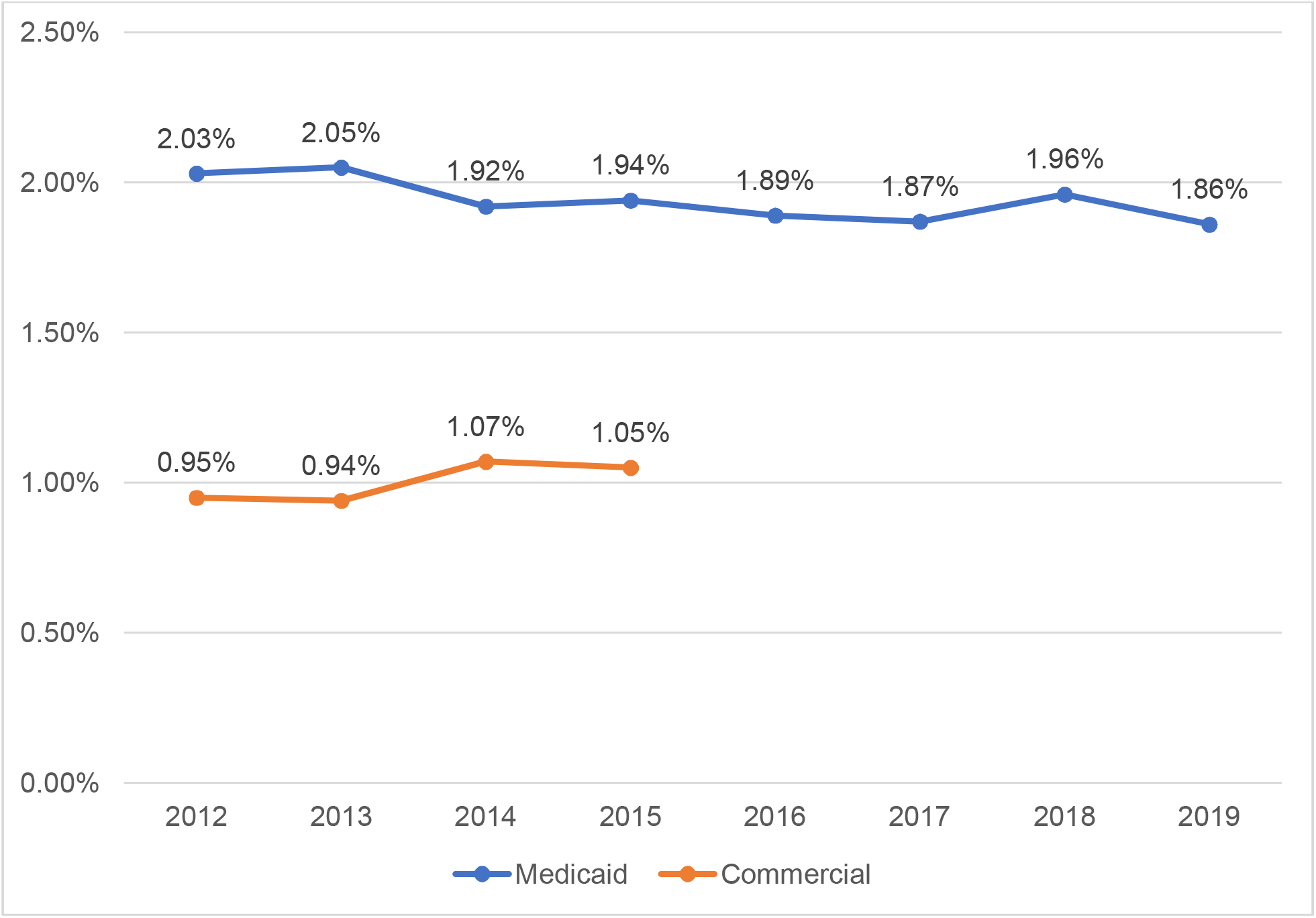
Oral/oropharyngeal cancer annual mortality rate, by cohort and year

### Cost of Oral Cancer

Overall, the total costs associated with the treatment of OC/OPC were higher for commercial insurance enrollees than for Medicaid enrollees by an average of $8.6 million between 2016 and 2019 (data not shown). When examining the cost of non-chemotherapy OC/OPC treatments, costs were higher for commercial enrollees across all three years by an average of $8.7 million as compared to those for Medicaid enrollees. However, the cost of chemotherapy was higher for Medicaid enrollees across these four years by an average of nearly $111,000, with Medicaid chemotherapy costs decreasing from $380,768 in 2016 to $189,236 in 2019.

Due to significant variability in cost per individual, both the mean and median cost per individual were calculated (Table 3). Mean and median commercial claims costs consistently were higher than for Medicaid claims. Between 2016 and 2019, Medicaid individual mean costs ranged from $8557.93 to $9502.78, while mean costs in commercial claims ranged from $21, 141.34 to $23,751.53. Median individual costs ranged from $583.72 to $661.51 per Medicaid enrollee and from $944.57 to $1,160.27 per commercial plan enrollee.

**Table 3:**
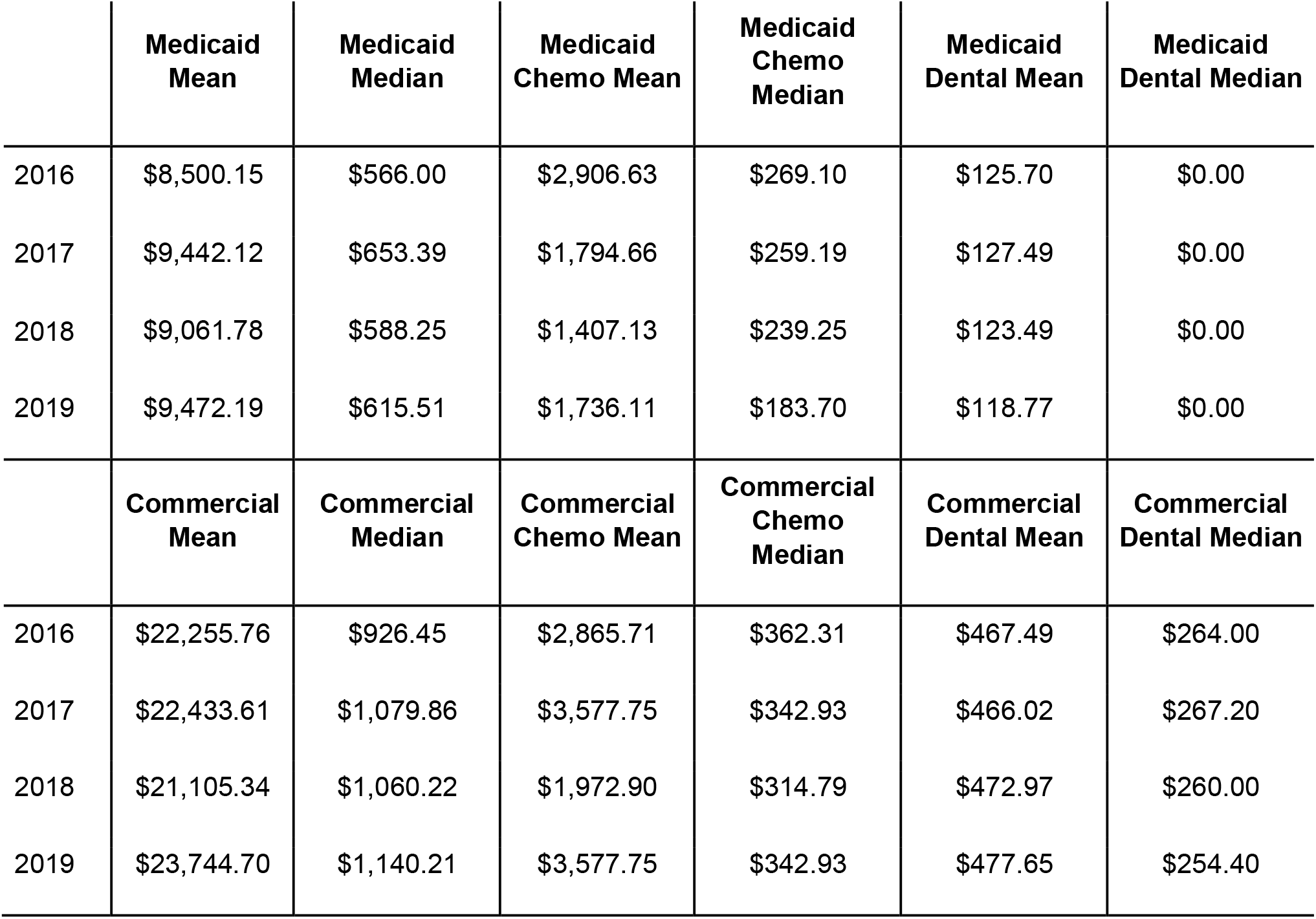
Costs of oral/oropharyngeal cancer in Medicaid and commercial insurance cohorts

## Discussion

Comparing large cohorts of adults covered by Medicaid or commercial insurance gives a more robust picture of the prevalence and incidence of OC/OPC in U.S. adults than would be achieved through an analysis of either group alone. Overall, the prevalence and incidence of OC/OPC were higher among Medicaid-enrolled adults than among those covered by commercial insurance across time, age groups, and sex. Uninsured individuals and those enrolled in Medicaid are less likely to have cancer screenings that may detect disease at a pre-malignant stage compared with individuals with private insurance,^(38-41)^ potentially resulting in higher levels of Medicaid enrollees being diagnosed with cancer than their privately insured counterparts. Further, individuals who are uninsured or are insured through Medicaid are more likely than individuals with private insurance to be diagnosed with advanced-stage cancers of various types.^(42-44)^ Because individuals enrolled in Medicaid must earn below certain income thresholds which are a percentage of the federal poverty level (Medicaid.gov), many may experience other environmental and behavioral factors associated with lower socioeconomic status and increased health risks, such as tobacco use, alcohol use, and lack of access to nutritious foods. ^(45)^

Similar to prior research,^(9)^ OC/OPC prevalence and incidence rates in our samples decreased over the course of six years (2012-18). We found a slightly higher prevalence of cancer in oropharyngeal sites compared to sites in the oral cavity, although we did not distinguish between specific sites in the oral and oropharyngeal regions. Given that predictors of OC/OPC in our sample included increased age and use of tobacco or alcohol, our results may be more reflective of non-HPV-related cases that have been declining over the last few decades. However, because there is not a clear consensus regarding which sites constitute oral versus oropharyngeal cancer,^(10)^ some readers may interpret our study’s data differently depending on how they define the anatomic sites included in each diagnosis.

In our study, individuals who visited a dentist within the previous year were less likely than those who did not to have an OC/OPC diagnosis. Regular dental visits provide frequent opportunities for dentists to detect OC/OPC at earlier stages.^(33, 34, 46, 47)^ Likely as a result, routine dental care is associated with improved survival from oral cancer,^(48, 49)^ and a greater likelihood of incident oral cancer is associated with infrequent dental care.0^(50, 51)^

In our sample, OC/OPC prevalence and incidence both increased with age, with the highest rates in the 60–64 age group in both the Medicaid and commercial insurance groups. That finding is consistent with those of Mahal and colleagues, who noted a unimodal peak in incidence between the ages of 60 to 64.^(52)^ Similarly, the median age of diagnosis of OC/OPC in the United States is 63 years, which corresponds with our findings.^(53)^ Interestingly, there has been a trend in the last few decades of an increase in OC/OPC incidence among younger individuals. Prior studies have found an increase in cancer of the tongue in adults under the age of 40.^(54, 55)^ As noted previously, this shift in incidence rates away from oral to more tongue and oropharyngeal sites has been attributed by some to an increase in HPV infections among younger populations, although the research in this area is mixed. There were noticeable increases between age groups in both prevalence and incidence beginning with individuals in their 40’s. For example, for Medicaid enrollees, there was a 3.6-fold increase in incidence (from 30.9 to 112.7) between adults aged 31–40 years and those aged 41–50 years, and then another 2.9-fold increase to those aged 51–60 years.

Consistent with prior research,^(1, 10, 12)^ the prevalence and incidence of OC/OPC in our sample was greater in males than females in both datasets. In a study comparing rates of oral cancer in Colorado to those of the U.S., Ernster and colleagues found that rates increased for males in both samples (36.6% in Colorado and 10.8% in the U.S.) between 1980-1991 and 1991-2000.^(28)^ Meanwhile, the rates of oral cancer for females decreased across the same period. The risk of death from oral cancer is higher for males than for females,^(56)^ and Tota and colleagues predicted a “substantial shift in burden [of oral cancer] to elderly white men” through 2029 based on increases in oral cancer in cohorts born before 1955.^(57)^

In our sample, OC/OPC prevalence was higher in White Medicaid enrollees compared with non-Hispanic Black or Hispanic enrollees or enrollees who reported an “other” racial or ethnic background. Racial or ethnic background data were not available in the commercial data. While cancer incidence has been declining since the mid-1980s overall, this decrease has been more gradual among White men; HPV-related cases have risen most dramatically in White men compared to other racial groups.^(58)^ Other research has noted an increase in oral cancer in White women under the age of 50 years.^(59)^ As noted by Boscolo-Rizzo and colleagues, “Patients with HPV-induced (oropharyngeal squamous cell carcinoma) are more likely to be middle-aged white men, non-smokers, non-drinkers or mild to moderate drinkers, with higher socioeconomic status…than subjects with HPV-unrelated (squamous cell carcinoma).”^(60)^ Conversely, a study by Ryerson and colleagues^(61)^ found the HPV-related incidence of oropharyngeal and oral cancers was highest in Black individuals. Further, Black individuals are more likely to be diagnosed with oral cancer at a younger age,^(62)^ have longer delays between diagnosis and treatment^(63)^ and have worse survival rates than their white counterparts.^(64)^

Mortality rates remained relatively consistent for Medicaid enrollees in our sample, ranging from 2.16% in 2012 to 2.12% in 2019, with a brief decrease to 1.94% in 2015. Similar rates were only available for commercially insured adults from 2012 through 2015 and showed an increase from 1.53% to 1.87% during that time. Rates from both groups were slightly lower than those from the National Cancer Institute’s Surveillance, Epidemiology, and End Results (SEER) program,^(53)^ which estimated the mortality rate from oral cavity and pharynx cancer as 2.5 per 100,000 each year based on 2014-2018 cases and deaths. Mortality rates were higher across all years for Medicaid enrollees compared to commercial enrollees, which is consistent with prior literature. Compared with private insurance recipients, Medicaid enrollees have worse OC/OPC mortality rates.^(65-68)^

Although the overall cost of OC/OPC treatment was higher for commercial enrollees in our sample, the cost for chemotherapy was higher for Medicaid enrollees. This likely reflects, at least in part, higher reimbursement rates for commercial plans compared to Medicaid. Jacobson and colleagues^(7)^ found that total annual health care spending for oral cancer patients was highest for patients with commercial insurance, followed by spending for those with Medicare and then Medicaid. In a study of quality of cancer care by insurance type in California, Parikh-Patel and colleagues^(69)^ found that individuals with Medicaid or dual eligibility for Medicare and Medicaid were significantly less likely than their privately insured peers to receive radiation and/or chemotherapy after diagnosis as recommended by the Commission on Cancer Quality Measures.

### Limitations

There are some limitations to the present study. These results come from claims data, which provide little information on factors such as stage at diagnosis, treatment decisions, or potential causes. Individuals over the age of 65 were not included in the analyses, as individuals who are dually eligible for Medicaid and Medicare would have most, if not all, of their cancer treatment charges covered by Medicare. Thus, analyses for those over the age of 65 would be underestimates. Because the median age of diagnosis for oral cancer in the United States is 63 years,^(53)^ that exclusion limits our ability to draw conclusions about a sizeable portion of the population with OC/OPC. Because data on racial or ethnic background were not available for the commercial insurance enrollees, it was not possible to determine potential disparities in that cohort. Finally, ICD-10 codes from medical claims were used to identify OC/OPC cases, which may provide a less precise accounting of cases than would medical records or similar sources.^(7)^

## Conclusion

This large-scale analysis of Medicaid and commercial claims data found an increased prevalence and incidence of OC/OPC and a higher mortality in Medicaid enrollees compared to their commercially insured counterparts. Having seen a dentist within the prior year was associated with a lower risk of having an OC/OPC diagnosis. As in prior studiesstudiesstudies, males and older adults were more likely to be diagnosed with OC/OPC than female and younger individuals. Non-Hispanic white Medicaid enrollees were more likely than non-Hispanic Black or Hispanic enrollees or those who identified as an “other” racial background to be diagnosed. Overall cancer treatment costs were higher for those with commercial insurance, although chemotherapy was more expensive for those enrolled in Medicaid. Risk factors for OC/OPC included male sex, older age, white race, tobacco or alcohol use, and comorbid immune disorders.

## Data Availability

All data produced in the present study are available upon reasonable request to the authors

## Appendix A: Prevalence* and incidence* of OC/OPC in Medicaid and commercial insurance groups, by year

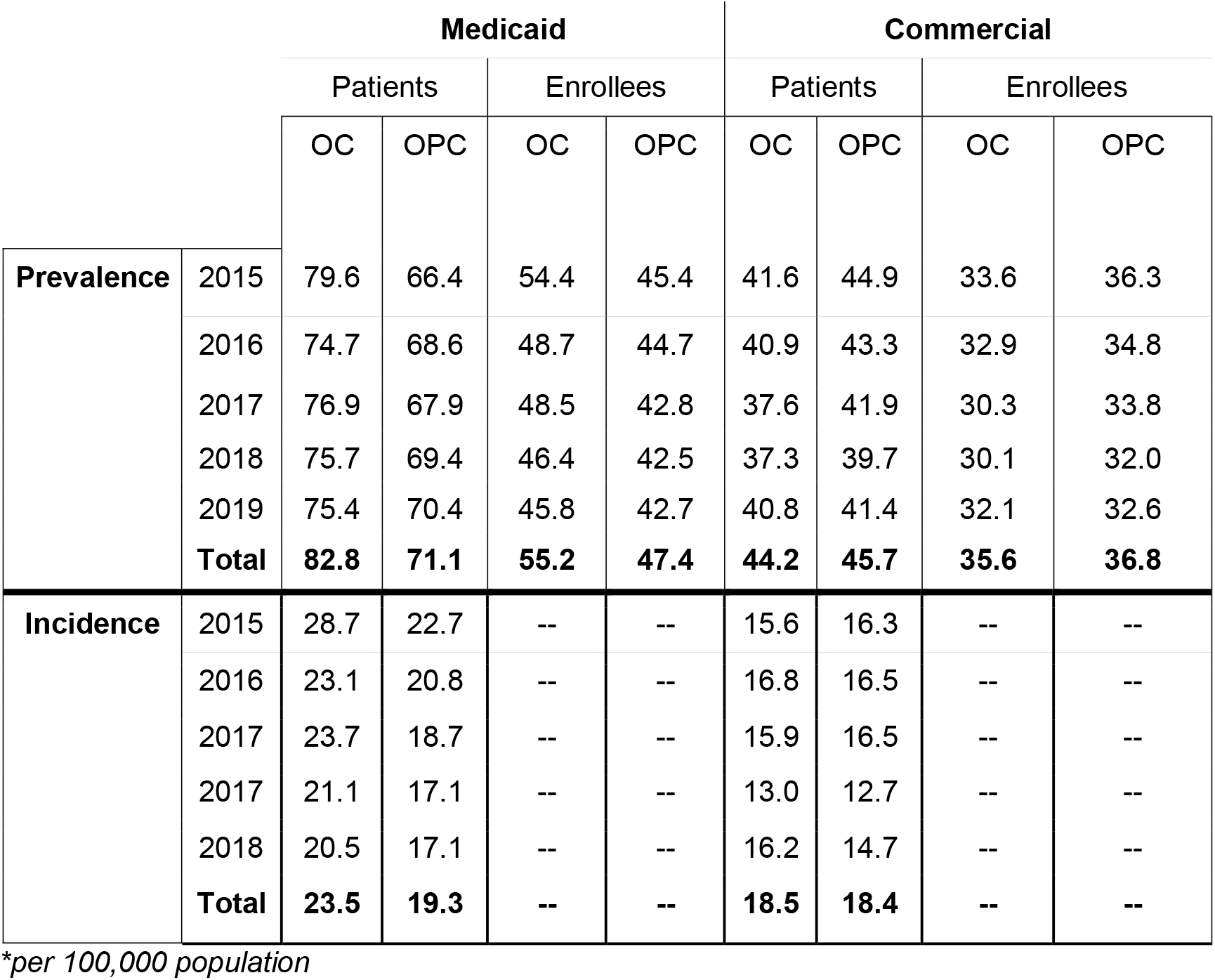

## Appendix B: Prevalence* of oral/oropharyngeal cancer among Medicaid and commercial insurance cohorts, by age group, 2012–2019 (combined)

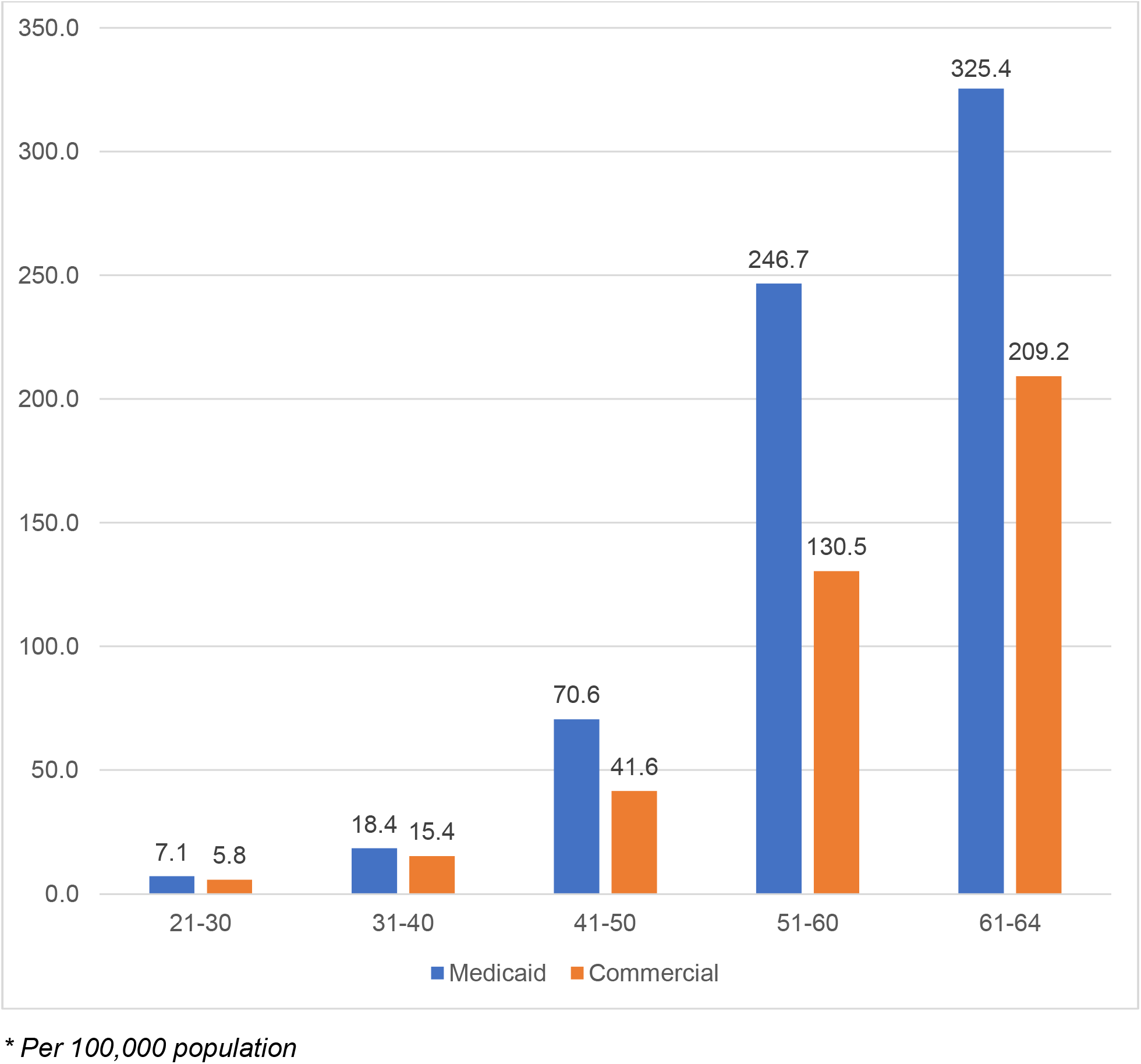

## Appendix C: Annual incidence* of oral/oropharyngeal cancer among Medicaid and commercial insurance cohorts, by age group

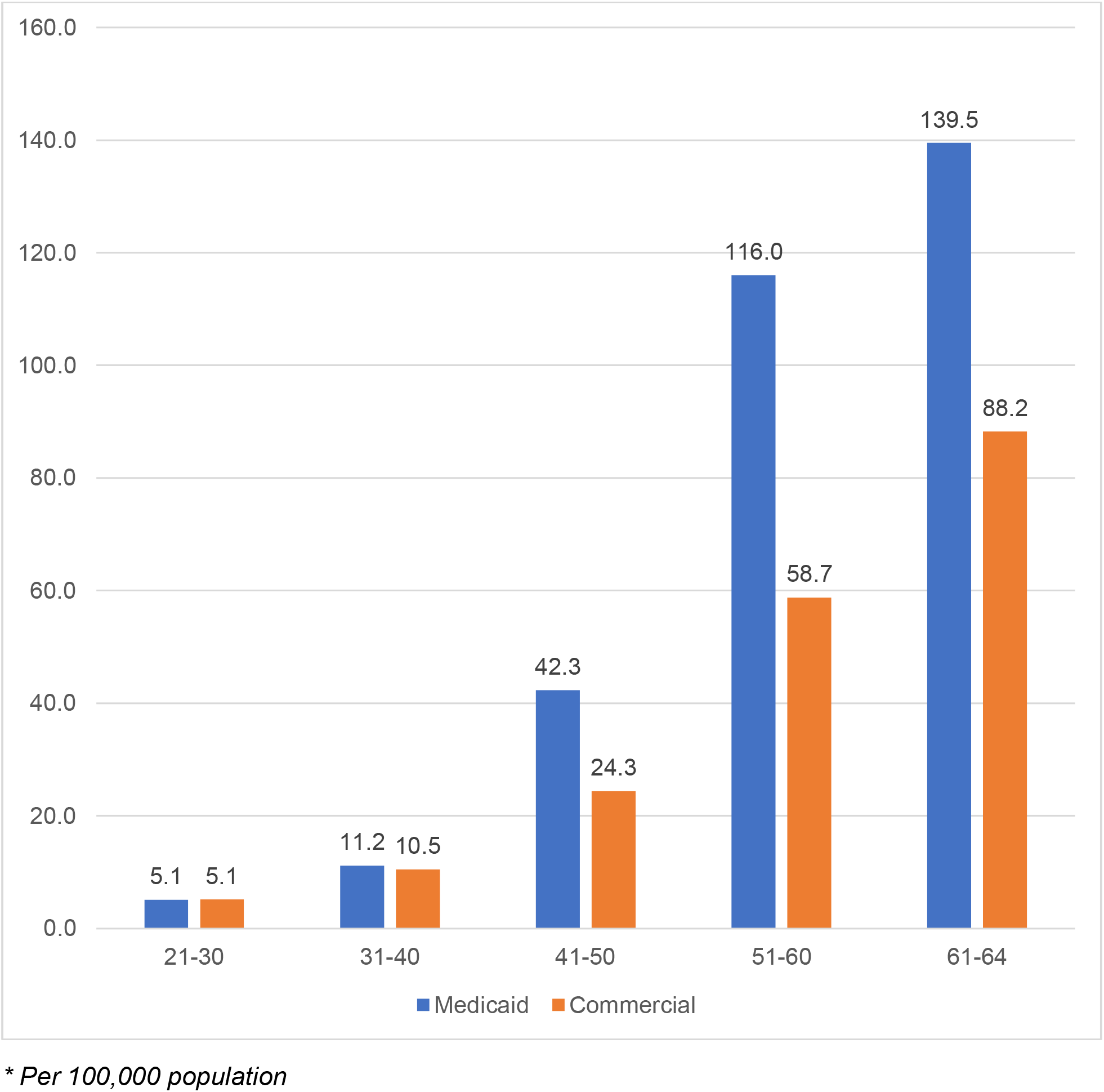

## References

1. Rivera C. Essentials of oral cancer. Int J Clin Exp Pathol. 2015;8(9):11884–94.

2. Sarode G, Maniyar N, Sarode SC, Jafer M, Patil S, Awan KH. Epidemiologic aspects of oral cancer. Dis Mon. 2020;66(12):100988.

3. Vigneswaran N, Williams MD. Epidemiologic trends in head and neck cancer and aids in diagnosis. Oral Maxillofac Surg Clin North Am. 2014;26(2):123–41.

4. Sung H, Ferlay J, Siegel RL, Laversanne M, Soerjomataram I, Jemal A, et al. Global Cancer Statistics 2020: GLOBOCAN Estimates of Incidence and Mortality Worldwide for 36 Cancers in 185 Countries. CA Cancer J Clin. 2021;71(3):209–49.

5. Bray F, Ferlay J, Soerjomataram I, Siegel RL, Torre LA, Jemal A. Global cancer statistics 2018: GLOBOCAN estimates of incidence and mortality worldwide for 36 cancers in 185 countries. CA Cancer J Clin. 2018;68(6):394–424.

6. Ellington TD, Henley SJ, Senkomago V, O’Neil ME, Wilson RJ, Singh S, et al. Trends in Incidence of Cancers of the Oral Cavity and Pharynx - United States 2007-2016. MMWR Morb Mortal Wkly Rep. 2020;69(15):433–8.

7. Jacobson JJ, Epstein JB, Eichmiller FC, Gibson TB, Carls GS, Vogtmann E, et al. The cost burden of oral, oral pharyngeal, and salivary gland cancers in three groups: commercial insurance, Medicare, and Medicaid. Head Neck Oncol. 2012;4:15.

8. American Cancer Society. Key Statistics for Oral Cavity and Oropharyngeal Cancers.: American Cancer Society; 2021 [Available from: https://www.cancer.org/cancer/oral-cavity-and-oropharyngeal-cancer/about/key-statistics.html.

9. Lingen MW, Abt E, Agrawal N, Chaturvedi AK, Cohen E, D’Souza G, et al. Evidence-based clinical practice guideline for the evaluation of potentially malignant disorders in the oral cavity: A report of the American Dental Association. J Am Dent Assoc. 2017;148(10):712–27 e10.

10. Conway DI, Purkayastha M, Chestnutt IG. The changing epidemiology of oral cancer: definitions, trends, and risk factors. Br Dent J. 2018;225(9):867–73.

11. Van Dyne EA, Henley SJ, Saraiya M, Thomas CC, Markowitz LE, Benard VB. Trends in Human Papillomavirus-Associated Cancers - United States, 1999-2015. MMWR Morb Mortal Wkly Rep. 2018;67(33):918–24.

12. Chi AC, Day TA, Neville BW. Oral cavity and oropharyngeal squamous cell carcinoma--an update. CA Cancer J Clin. 2015;65(5):401–21.

13. Walvik L, Svensson AB, Friborg J, Lajer CB. The association between human papillomavirus and oropharyngeal squamous cell Carcinoma: Reviewed according to the Bradford Hill criteria for causality. Oral Oncol. 2016;63:61–5.

14. Chaturvedi AK, Engels EA, Anderson WF, Gillison ML. Incidence trends for human papillomavirus-related and -unrelated oral squamous cell carcinomas in the United States. J Clin Oncol. 2008;26(4):612–9.

15. Chaturvedi AK, Engels EA, Pfeiffer RM, Hernandez BY, Xiao W, Kim E, et al. Human papillomavirus and rising oropharyngeal cancer incidence in the United States. J Clin Oncol. 2011;29(32):4294–301.

16. D’Souza G, Westra WH, Wang SJ, van Zante A, Wentz A, Kluz N, et al. Differences in the Prevalence of Human Papillomavirus (HPV) in Head and Neck Squamous Cell Cancers by Sex, Race, Anatomic Tumor Site, and HPV Detection Method. JAMA Oncol. 2017;3(2):169–77.

17. Sturgis EM, Cinciripini PM. Trends in head and neck cancer incidence in relation to smoking prevalence: an emerging epidemic of human papillomavirus-associated cancers? Cancer. 2007;110(7):1429–35.

18. Kreimer AR, Clifford GM, Boyle P, Franceschi S. Human papillomavirus types in head and neck squamous cell carcinomas worldwide: a systematic review. Cancer Epidemiol Biomarkers Prev. 2005;14(2):467–75.

19. Patel SC, Carpenter WR, Tyree S, Couch ME, Weissler M, Hackman T, et al. Increasing incidence of oral tongue squamous cell carcinoma in young white women, age 18 to 44 years. J Clin Oncol. 2011;29(11):1488–94.

20. Tota JE, Anderson WF, Coffey C, Califano J, Cozen W, Ferris RL, et al. Rising incidence of oral tongue cancer among white men and women in the United States, 1973-2012. Oral Oncol. 2017;67:146–52.

21. Toporcov TN, Znaor A, Zhang ZF, Yu GP, Winn DM, Wei Q, et al. Risk factors for head and neck cancer in young adults: a pooled analysis in the INHANCE consortium. Int J Epidemiol. 2015;44(1):169–85.

22. Muller S, Pan Y, Li R, Chi AC. Changing trends in oral squamous cell carcinoma with particular reference to young patients: 1971-2006. The Emory University experience. Head Neck Pathol. 2008;2(2):60–6.

23. Hernandez DJ, Alam B, Kemnade JO, Huang AT, Chen AC, Sandulache VC. Consistent multimodality approach to oral cavity and high-risk oropharyngeal cancer in veterans. Am J Otolaryngol. 2021;42(6):103166.

24. Janz TA, Momin SR, Sterba KR, Kato MG, Armeson KE, Day TA. Comparison of psychosocial factors over time among HPV+ oropharyngeal cancer and tobacco-related oral cavity cancer patients. Am J Otolaryngol. 2019;40(1):40–5.

25. Warnakulasuriya S. Living with oral cancer: epidemiology with particular reference to prevalence and life-style changes that influence survival. Oral Oncol. 2010;46(6):407–10.

26. Epstein JB, Cabay RJ, Glick M. Oral malignancies in HIV disease: changes in disease presentation, increasing understanding of molecular pathogenesis, and current management. Oral Surg Oral Med Oral Pathol Oral Radiol Endod. 2005;100(5):571–8.

27. Chaitanya NC, Allam NS, Gandhi Babu DB, Waghray S, Badam RK, Lavanya R. Systematic meta-analysis on association of human papilloma virus and oral cancer. J Cancer Res Ther. 2016;12(2):969–74.

28. Ernster JA, Sciotto CG, O’Brien MM, Finch JL, Robinson LJ, Willson T, et al. Rising incidence of oropharyngeal cancer and the role of oncogenic human papilloma virus. Laryngoscope. 2007;117(12):2115–28.

29. Lafaurie GI, Perdomo SJ, Buenahora MR, Amaya S, Diaz-Baez D. Human papilloma virus: An etiological and prognostic factor for oral cancer? J Investig Clin Dent. 2018;9(2):e12313.

30. American Dental Association. ADA expands policy on oral cancer detection to include oropharyngeal cancer 2019 [Available from: https://www.ada.org/en/publications/ada-news/2019-archive/september/ada-expands-policy-on-oral-cancer-detection-to-include-oropharyngeal-cancer#.

31. Mignogna MD, Fedele S. Oral cancer screening: 5 minutes to save a life. Lancet. 2005;365(9475):1905–6.

32. Sankaranarayanan R, Ramadas K, Thomas G, Muwonge R, Thara S, Mathew B, et al. Effect of screening on oral cancer mortality in Kerala, India: a cluster-randomised controlled trial. Lancet. 2005;365(9475):1927–33.

33. Langevin SM, Michaud DS, Eliot M, Peters ES, McClean MD, Kelsey KT. Regular dental visits are associated with earlier stage at diagnosis for oral and pharyngeal cancer. Cancer Causes Control. 2012;23(11):1821–9.

34. Watson JM, Logan HL, Tomar SL, Sandow P. Factors associated with early-stage diagnosis of oral and pharyngeal cancer. Community Dent Oral Epidemiol. 2009;37(4):333–41.

35. Tiwari T, Tranby E, Thakkar-Samtani M, Frantsve-Hawley J. Determinants of Tooth Loss in a Medicaid Adult Population. JDR Clin Trans Res. 2021:23800844211022277.

36. Dye BA, Tan S, Smith V, Lewis BG, Barker LK, Thornton-Evans G, et al. Trends in oral health status: United States, 1988-1994 and 1999-2004. Vital Health Stat 11. 2007(248):1–92.

37. Eke PI, Dye BA, Wei L, Thornton-Evans GO, Genco RJ, Cdc Periodontal Disease Surveillance workgroup: James Beck GDRP. Prevalence of periodontitis in adults in the United States: 2009 and 2010. J Dent Res. 2012;91(10):914–20.

38. Ayanian JZ, Weissman JS, Schneider EC, Ginsburg JA, Zaslavsky AM. Unmet health needs of uninsured adults in the United States. JAMA. 2000;284(16):2061–9.

39. Ioannou GN, Chapko MK, Dominitz JA. Predictors of colorectal cancer screening participation in the United States. Am J Gastroenterol. 2003;98(9):2082–91.

40. Potosky AL, Breen N, Graubard BI, Parsons PE. The association between health care coverage and the use of cancer screening tests. Results from the 1992 National Health Interview Survey. Med Care. 1998;36(3):257–70.

41. Ward E, Halpern M, Schrag N, Cokkinides V, DeSantis C, Bandi P, et al. Association of insurance with cancer care utilization and outcomes. CA Cancer J Clin. 2008;58(1):9–31.

42. Kwok J, Langevin SM, Argiris A, Grandis JR, Gooding WE, Taioli E. The impact of health insurance status on the survival of patients with head and neck cancer. Cancer. 2010;116(2):476–85.

43. Rapado D, Chowdhari S, Gu C, Varella M, Castro G, Rodriguez de la Vega P, et al. Impact of Insurance Status on Diagnostic Stage in Hodgkin’s Lymphoma in the United States: Implications for Detection and Outcomes. Cureus. 2020;12(11):e11600.

44. Zhao J, Han X, Nogueira L, Jemal A, Halpern MT, Yabroff KR. Health insurance status and cancer stage at diagnosis and survival in the United States. Journal of Clinical Oncology. 2020;38(15_suppl):7026–.

45. Ward E, Jemal A, Cokkinides V, Singh GK, Cardinez C, Ghafoor A, et al. Cancer disparities by race/ethnicity and socioeconomic status. CA Cancer J Clin. 2004;54(2):78–93.

46. Elwood JM, Gallagher RP. Factors influencing early diagnosis of cancer of the oral cavity. CMAJ. 1985;133(7):651–6.

47. Groome PA, Rohland SL, Hall SF, Irish J, Mackillop WJ, O’Sullivan B. A population-based study of factors associated with early versus late stage oral cavity cancer diagnoses. Oral Oncol. 2011;47(7):642–7.

48. Farquhar DR, Divaris K, Mazul AL, Weissler MC, Zevallos JP, Olshan AF. Poor oral health affects survival in head and neck cancer. Oral Oncol. 2017;73:111–7.

49. Haynes DA, Vanison CC, Gillespie MB. The Impact of Dental Care in Head and Neck Cancer Outcomes: A Systematic Review and Meta-Analysis. Laryngoscope. 2021.

50. Ali K, Kay EJ. Is there an association between past dental visits and the incidence of cancers of the head and neck (HN), upper aerodigestive tract (UADT), and oral cavity? Evid Based Dent. 2019;20(2):37–8.

51. Gupta B, Kumar N, Johnson NW. Evidence of past dental visits and incidence of head and neck cancers: a systematic review and meta-analysis. Syst Rev. 2019;8(1):43.

52. Mahal BA, Catalano PJ, Haddad RI, Hanna GJ, Kass JI, Schoenfeld JD, et al. Incidence and Demographic Burden of HPV-Associated Oropharyngeal Head and Neck Cancers in the United States. Cancer Epidemiol Biomarkers Prev. 2019;28(10):1660–7.

53. Howlader NN, A.M.; Krapcho, M.; Miller, D.; Brest, A.; Yu, M.; Ruhl, J.; Tatalovich, Z.; Mariotto, A.; Lewis, D.R.; Chen, H.S.; Feuer, E.J.; Cronin, K.A. (eds). SEER Cancer Statistics Review, 1975-2017 Bethesda, MD: National Cancer Institute; 2020 [

54. Schantz SP, Yu GP. Head and neck cancer incidence trends in young Americans, 1973-1997, with a special analysis for tongue cancer. Arch Otolaryngol Head Neck Surg. 2002;128(3):268–74.

55. Shiboski CH, Schmidt BL, Jordan RC. Tongue and tonsil carcinoma: increasing trends in the U.S. population ages 20-44 years. Cancer. 2005;103(9):1843–9.

56. Fakhry C, Westra WH, Wang SJ, van Zante A, Zhang Y, Rettig E, et al. The prognostic role of sex, race, and human papillomavirus in oropharyngeal and nonoropharyngeal head and neck squamous cell cancer. Cancer. 2017;123(9):1566–75.

57. Tota JE, Gillison ML, Katki HA, Kahle L, Pickard RK, Xiao W, et al. Development and validation of an individualized risk prediction model for oropharynx cancer in the US population. Cancer. 2019;125(24):4407–16.

58. Brown LM, Check DP, Devesa SS. Oropharyngeal cancer incidence trends: diminishing racial disparities. Cancer Causes Control. 2011;22(5):753–63.

59. Joseph LJ, Goodman M, Higgins K, Pilai R, Ramalingam SS, Magliocca K, et al. Racial disparities in squamous cell carcinoma of the oral tongue among women: a SEER data analysis. Oral Oncol. 2015;51(6):586–92.

60. Boscolo-Rizzo P, Del Mistro A, Bussu F, Lupato V, Baboci L, Almadori G, et al. New insights into human papillomavirus-associated head and neck squamous cell carcinoma. Acta Otorhinolaryngol Ital. 2013;33(2):77–87.

61. Ryerson AB, Peters ES, Coughlin SS, Chen VW, Gillison ML, Reichman ME, et al. Burden of potentially human papillomavirus-associated cancers of the oropharynx and oral cavity in the US, 1998-2003. Cancer. 2008;113(10 Suppl):2901–9.

62. Mukherjee A, Idigo AJ, Ye Y, Wiener HW, Paluri R, Nabell LM, et al. Geographical and Racial Disparities in Head and Neck Cancer Diagnosis in South-Eastern United States: Using Real-World Electronic Medical Records Data. Health Equity. 2020;4(1):43–51.

63. Naghavi AO, Echevarria MI, Strom TJ, Abuodeh YA, Ahmed KA, Venkat PS, et al. Treatment delays, race, and outcomes in head and neck cancer. Cancer Epidemiol. 2016;45:18–25.

64. Clarke JA, Despotis AM, Ramirez RJ, Zevallos JP, Mazul AL. Head and Neck Cancer Survival Disparities by Race and Rural-Urban Context. Cancer Epidemiol Biomarkers Prev. 2020;29(10):1955–61.

65. Agarwal P, Agrawal RR, Jones EA, Devaiah AK. Social Determinants of Health and Oral Cavity Cancer Treatment and Survival: A Competing Risk Analysis. Laryngoscope. 2020;130(9):2160–5.

66. Challapalli SD, Simpson MC, Adjei Boakye E, Pannu JS, Costa DJ, Osazuwa-Peters N. Head and Neck Squamous Cell Carcinoma in Adolescents and Young Adults: Survivorship Patterns and Disparities. J Adolesc Young Adult Oncol. 2018;7(4):472–9.

67. Gaubatz ME, Bukatko AR, Simpson MC, Polednik KM, Adjei Boakye E, Varvares MA, et al. Racial and socioeconomic disparities associated with 90-day mortality among patients with head and neck cancer in the United States. Oral Oncol. 2019;89:95–101.

68. Panth N, Simpson MC, Sethi RKV, Varvares MA, Osazuwa-Peters N. Insurance status, stage of presentation, and survival among female patients with head and neck cancer. Laryngoscope. 2020;130(2):385–91.

69. Parikh-Patel A, Morris CR, Kizer KW. Disparities in quality of cancer care: The role of health insurance and population demographics. Medicine (Baltimore). 2017;96(50):e9125.

